# Perinatal SSRI Exposure Impacts Innate Fear Circuit Activation and Behavior in Mice and Humans

**DOI:** 10.1101/2023.03.01.23286641

**Authors:** Giulia Zanni, Milenna Van Dijk, Martha Caffrey Cagliostro, Gregory S. Stevens, Nicolò Pini, Ariel L. Rose, Alexander L. Kesin, Claudia Lugo-Candelas, Priscila Dib Goncalves, Alexandra S. MacKay, Praveen Kulkarni, Craig F. Ferris, Myrna M. Weissman, Ardesheer Talati, Mark S. Ansorge, Jay A. Gingrich

## Abstract

Serotonin shapes brain structure and function during early development across phylogenetically diverse species. In mice and humans, perinatal SSRI exposure produces brain alterations and increases anxiety/depression-related behaviors in the offspring. It remains unclear whether shared brain circuit changes underlie the behavioral impact of perinatal SSRIs across species. We examine how developmental SSRI-exposure in mice and humans changes fear-related brain activation and behavior. SSRI-administered mice showed increased defense responses to a predator odor that were associated with stronger fMRI-based fear circuit activation when compared to saline controls. Similarly, human adolescents exposed to SSRIs *in utero* showed greater activation of fear brain structures and exhibited higher anxiety and depressive symptoms than unexposed adolescents. Perinatal SSRI enhances innate fear-related responses and fear brain circuit activation that are conserved across species.

**One Sentence Summary:** Since SSRI use in pregnancy is common, we determined the effects of altered serotonin signaling during development in mice and humans.

## Main Text

Maternal anxiety and depression during pregnancy are associated with adverse offspring outcomes ranging from low birth weight to increased risk for psychopathology including anxiety and depression^1–3^. Such maternal symptoms are typically treated with selective serotonin reuptake inhibitors (SSRIs). Currently taken by ∼6% of pregnant mothers^4^, their use is increasing^5^ due to treatment recommendations from advisory agencies. Although therapeutically effective for the mother, SSRIs cross the placenta into the fetal brain and increase fetal serotonin levels^6^. Serotonin (5-hydroxytryptamine [5-HT]) signaling is crucial to fetal brain development and plays a role in processes such as cell proliferation, neuronal differentiation, synaptogenesis, and neuronal migration^7–9^. Perinatal increases in 5-HT signaling lead to altered structure and function throughout the brain in mice^10,11^. Although 5-HT is one of the most phylogenetically-conserved neurotransmitters, found in the central nervous system from mollusks to primates^12–14^, it remains unclear whether SSRI exposure during early brain development exerts in humans the same effects seen in rodents.

While SSRI-exposure *in utero* may have few detrimental effects on human offspring in early infancy (but see^15–17^), some studies report cognitive and behavioral abnormalities in later childhood^18,19^. Rodent^20,21^ and some epidemiological^22,23^ studies suggest that SSRI use in pregnancy may be associated with depression and other psychiatric disorders in the offspring starting in early adolescence^23,24^, but human studies have not always been consistent^25,26^. Rodent experiments demonstrate a clear link between developmental exposure to SSRIs and later behavioral outcomes, and avoid most of the confounds of large human studies. However, there has not been a study comparing the circuit-level activation of regions related to anxiety and depression across species as a consequence of early-life SSRI exposure.

Human neuroimaging studies show an association between prenatal SSRI exposure and neural correlates in newborns and infants. Specifically, in neonates prenatal SSRI exposure is associated with increased amygdala and insula volume^27^, decreased microstructure in fronto-thalamic and fronto-fugal tracts amongst others^28^, and alterations in signal processing measured by resting state fMRI^29^ and EEG^30,31^. However, such consequences of prenatal SSRI use have not been investigated in older offspring that are more developed and more proximate to possible behavioral sequelae^32^.

In mice SSRI administration during postnatal days (PND) 2-11 (corresponding to the third trimester of human pregnancy) results in several anxiety-related behavioral impairments, such as increased conditioned fear response and reductions in cued fear extinction linked to amygdala dysfunction^20,33–36^. Because the amygdala is also part of a neural network mediating behavioral responses to innate fear cues (e.g. predator odors in rodents or fearful faces in humans)^37,38,39^, such responses might also be vulnerable to early-life SSRI exposure.

Here, we adopt a cross-species approach that harnesses controlled experimental conditions, and tests clinical implications within a single study framework. We compare the innate fear responses in mice and human adolescents exposed to SSRIs (PND2-11 for mice; *in utero* for human adolescents). Specifically, we examine the behavioral effects of perinatal SSRI exposure on innate fear responses to predator odor in mice. To allow comparison to human subjects, we use functional magnetic resonance imaging (fMRI) in awake mice during predator odor presentation and compare the differential brain activation in fluoxetine-versus vehicle-treated adult animals. For human participants, we used fMRI to assess BOLD response to fearful compared to neutral faces in adolescent children exposed and unexposed to prenatal SSRIs. Our results suggest that early-life exposure to SSRIs has similar effects on innate fear circuitry across phylogeny and has public health implications for the broad use of SSRI medications during pregnancy.

### PND2-11 fluoxetine increases freezing and crouching to a predator but not neutral odor in adult mice

Previous work identified PND2-11 as a sensitive period for 5-HT levels in mice that impacts conditioned fear responses and anxiety in the adult^20,33^. To increase translatability and establish fMRI compatibility, here we studied the effects of PND2-11 fluoxetine (PNFLX) on innate fear responses to a predator odor (mountain lion urine) (**Figure 1A**). Three-way ANOVA analysis of freezing during habituation and predator odor exposure revealed an interaction between treatment and predator odor (F _4,48_=0.4866, *p=0.0422), time and predator odor (F _4,48_=4.651, **p=0.0030), and a main effect of time (F _4,48_=4.851, *p=0.0023) (**Figure 1B)**. Post-hoc analysis showed that over the course of the predator odor test PNFLX animals had significantly higher freezing levels when compared to postnatal saline (PNSAL) animals, peaking 3 minutes into the test (**p=0.0095). A similar pattern of freezing was observed in adolescent PNFLX mice compared to PNSAL mice (**Figure 1 Supplementary**). For latency to freeze as percent of habituation (**Figure 1C**) we found a treatment x predator odor interaction (F _1,10_=5.416, *p=0.0422), a predator odor effect (F _1,10_=44.46, ****p<0.0001) as well as a treatment effect (F _1,10_=5.417, *p=0.0422). Post-hoc analysis revealed that PNFLX animals had a higher percent reduction in latency to freeze compared to PNSAL animals during the predator odor test (**p=0.0073). PNSAL animals had an average 43% reduction in latency to freezing during the predator odor test (*p=0.0236), while PNFLX animals had a 78% reduction in latency to freezing during the predator odor test compared to habituation (***p=0.0002). These data demonstrate that both groups of animals responded to the predator odor with freezing but the behavioral reaction is exacerbated in PNFLX mice. Three-way ANOVA analysis of total distance traveled revealed no treatment or treatment interaction significances, demonstrating that general exploration behavior during habituation and predator odor presentation is not different between PNSAL and PNFLX animals (Predator odor test F _1,12_=8.844, ***p=0.0116, Time F _4,48_=10.90, ****p<0.0001) (**Figure 1D**). To gain further insight into the behavioral response to predator odor, we used a deep learning algorithm that automates animal pose estimation to assess defensive crouching behavior with body perimeter as a proxy (**Figure 1E**). Plotting average perimeter against total freezing of all mice during both habituation and predator odor testing, we found a significant negative correlation between these two independent variables (*p=0.01007, r^2^=0.1908), suggesting that a smaller perimeter reflects defensive postures (**Figure 1F**). Because defensive responses adapt over time^40^, we investigated how perimeter changed over 5 minutes of predator odor exposure as percent of habituation. Two-way ANOVA analysis identified a predator odor x treatment interaction (F _4,51_=2.858, *p=0.0326), no effect of treatment (F _1,51_=0.9200, p=0.3420), and no effect of predator odor (F _4,51_=0.7792, p=0.5439). Post-hoc analysis showed that by the second minute of the test, PNFLX animals had significantly lower perimeter compared to PNSAL (*p=0.0136) (**Figure 1G**), again demonstrating an exaggerated and faster behavioral response to the predator odor.

**Figure 1.**
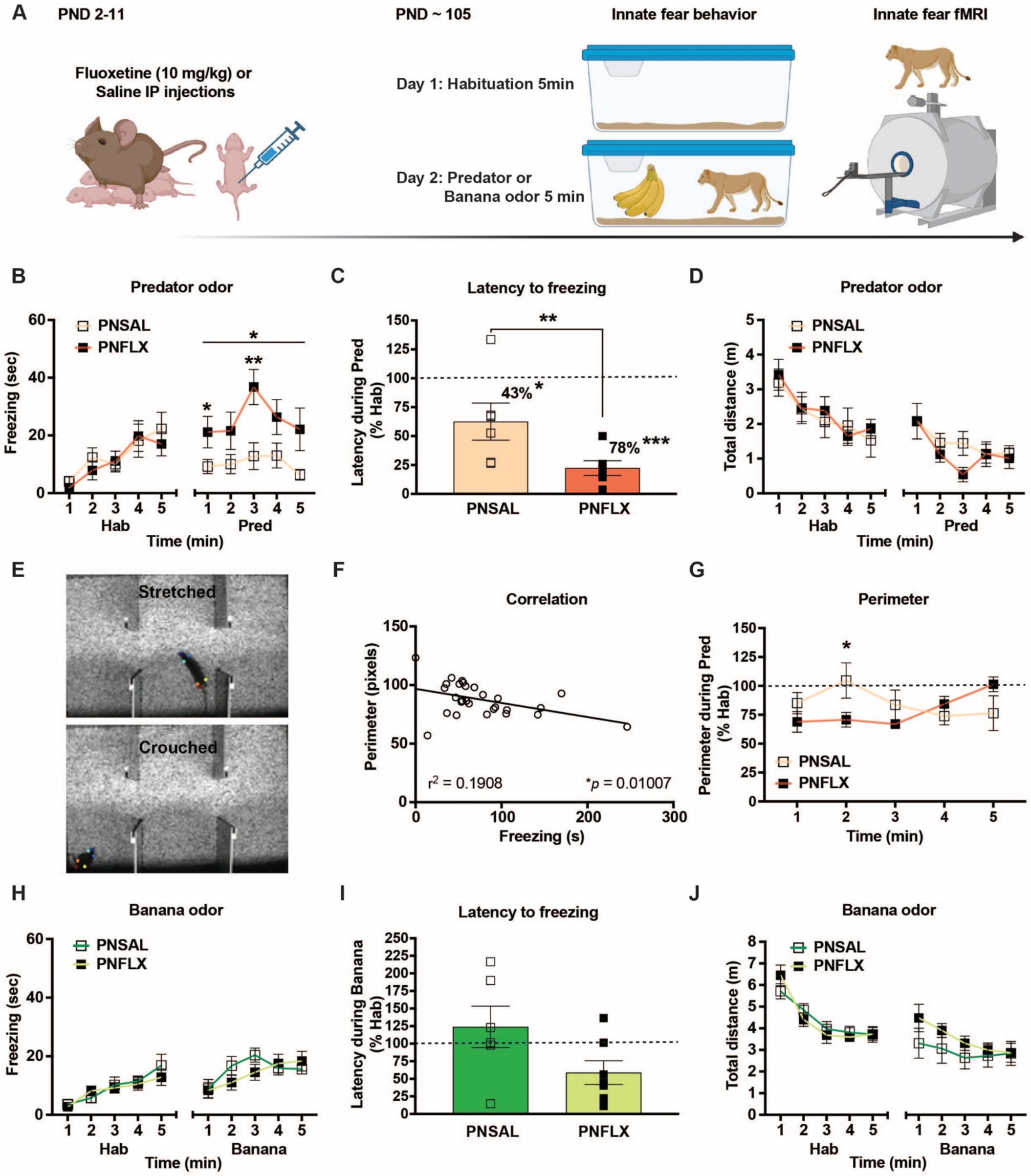
Postnatal fluoxetine (PND2-11) increases freezing and crouching to a predator but not neutral odor in adult mice. **A**) Timeline of the adult mouse study. **B**) Freezing over the 5 minutes of habituation and predator odor test. **C**) Percent reduction in latency to freeze. **D**) Total distance over time during habituation and predator odor test. **E**) Labeling of different animal body parts: nose (dark blue), tail base (electric blue), tail end (light blue), right ear (yellow), left ear (green), right hip (red), and left hip (orange). Top picture shows animal stretching and the bottom picture shows an animal crouching in different zones of the chamber (safe, neutral, and predator). **F**) Correlation between freezing and normalized perimeter. **G**) Normalized body perimeter to habituation baseline over the 5 minutes of predator odor test. **H**) Time course of freezing over the 5 minutes of habituation and banana odor test. **I**) Percent reduction in latency to freeze. **J**) Total distance over time during habituation and banana (neutral) odor. PNSAL N=5-7, PNFLX N=5-7. \**p<0*.*05*; \*\**p<0*.*01*; \*\*\**p<0*.*001*

To address the specificity of the immobility response to the predator odor, we examined the fear reaction to a neutral banana odor. Three-way ANOVA analysis of freezing over the 5 minutes of banana odor exposure showed no test x treatment x time interaction (F _4,48_=1.647, p=0.1780), nor effect of treatment (F _1,12_=0.4611, p = 0.5100). We found an effect of Test (F _1,12_=28.17, ***p=0.0002) and an effect of Time (F _4,48_=12.51, ****p<0.0001) but no interaction with treatment, suggesting that all animals irrespective of treatment are more immobile during the neutral odor exposure (**Figure 1H**). Analysis of the percent reduction in latency to freeze revealed that there was no test x treatment x time interaction (F _1,12_=1.838, p=0.2002), no effect of test (F _1,12_=1.005, p=0.3359), nor effect of treatment (F _1,12_=1.838, p=0.2002) (**Figure 1I**). During habituation PNSAL and PNFLX treated animals covered a similar distance (three-way ANOVA treatment F _1,12_=0.5583, p=0.4693), however during the banana odor test both PNSAL and PNFLX animals traveled less distance compared to the habituation period (Test F _1,12_=10.27, **p=0.0076, Time F _4,48_=40.05, ****p<0.0001) showing that all animals decreased their exploration during neutral odor (**Figure 1J**).

### PND2-11 fluoxetine increases activity in fear circuits in response to mountain lion predator-odor

To address which brain regions are contributing to the SSRI-induced heightened innate fear response in mice we used functional magnetic resonance imaging (fMRI) to compare activation in response to predator odor in PNFLX mice and PNSAL controls. To elicit fear in awake mice we exposed them to a predator odor in a holding system to which they were previously habituated and performed whole-brain imaging (**Figure 2A**). The fMRI scans were registered to and analyzed using a 3-dimensional MRI mouse atlas with 134 bilateral segmented and annotated brain areas. The atlas was used to construct the integrated neural circuitry comprising the key brain areas involved in the perception of fear as described in the rodent literature, which we hypothesized would be regions involved in the task and affected by SSRI exposure (**Figure 2B**). **Figure 2C** and the table in **Figure 2D** show the brain areas with a significant increase (α<0.05) in positive BOLD volume of activation in response to predator urine odor in awake PNFLX mice compared to PNSAL mice (false discovery rate p=0.0401) in whole-brain analysis. These areas are ranked in order of their significance and include several areas that make up the fear neural circuit e.g. central and medial amygdala, periaqueductal gray, anterior cingulate cortex, in addition to areas that comprise the ascending reticular activating system involved in arousal, e.g. pedunculopontine tegmental area, medullary reticular area, and mesencephalic reticular formation. The brain regions affected are highly conserved and include among others amygdala, hypothalamus, putamen, periaqueductal gray, and dorsal raphe. These are known to be key regulatory structures of emotional states. Overall this suggests that PND2-11 fluoxetine treatment leads to hyperactivation of the fear brain circuit in adult mice.

**Figure 2.**
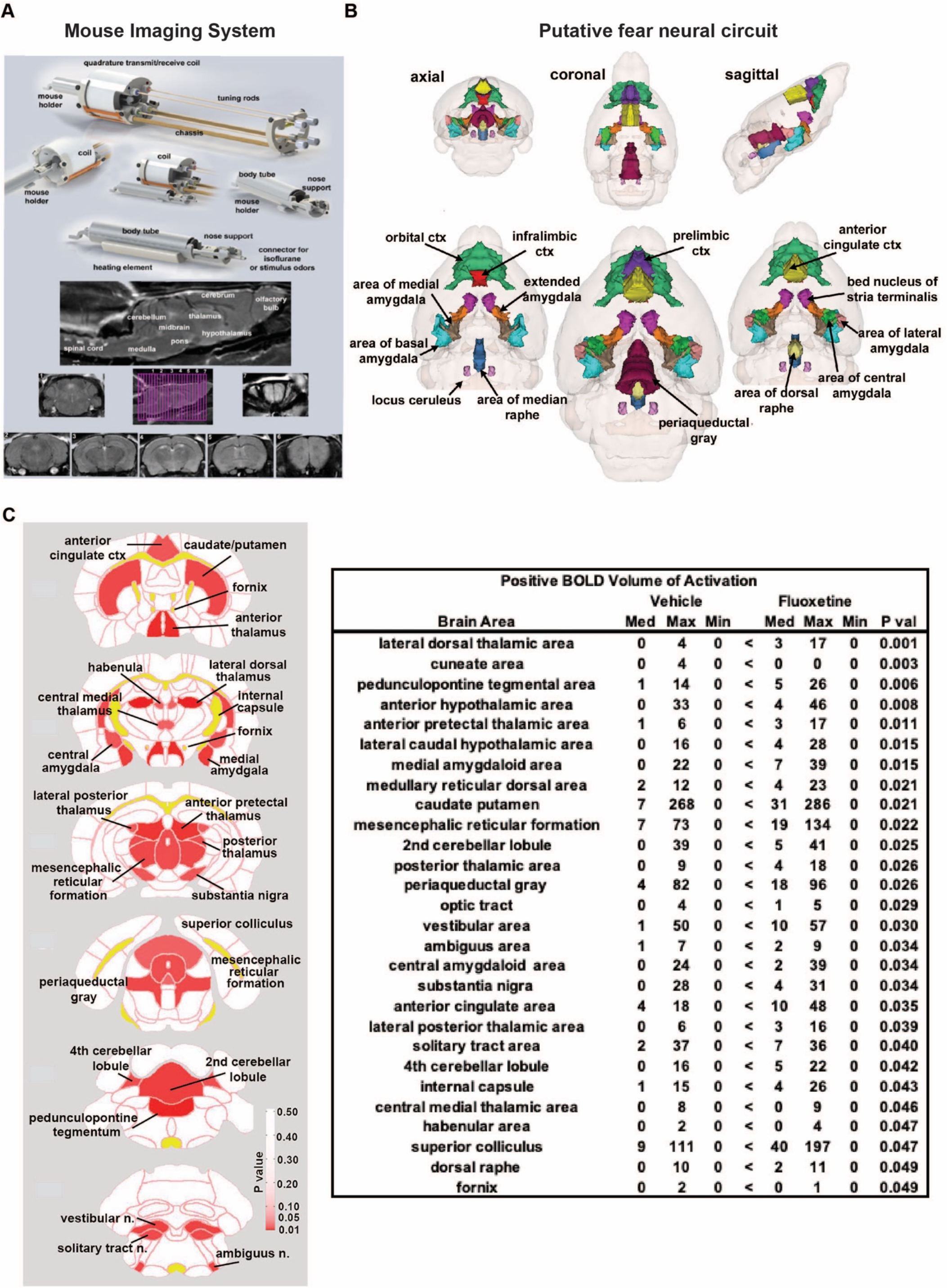
Postnatal fluoxetine (PND2-11) increases innate fear circuit activation. **A**) Shown are the different components of the mouse imaging system. Below are sagittal and axial views of an awake mouse brain. Note the linearity along the Z-axis. The axial images taken from a 22-slice RARE sequence (0.6mm thickness) demonstrate complete brain coverage from the olfactory bulbs to the brainstem. **B**) fMRI scans were aligned to a 3-dimensional MRI mouse brain atlas, and brain areas that comprise the fear neural circuit are highlighted. **C**) Statistical map displaying relative positive BOLD signal in response to predator urine odor in awake PNFLX compared to PNSAL mice. PNSAL N=18, PNFLX N=26. **D**) List of significant brain areas, such as thalamus, putamen, brainstem and cerebellum, central amygdala, substantia nigra, periaqueductal gray, raphe, and habenula ranked in order of their significance in red for change in positive BOLD volume of activation (number of voxels) in PNFLX compared to PNSAL (false discovery rate p=0.0401).

### Translation to human adolescent children from the ABCD study

In the rodents, fluoxetine was administered at PND2-11, corresponding to the third trimester *in utero* in humans. Since behavioral consequences of prenatal SSRIs have been observed in adolescence, we leveraged data from the Adolescent Brain and Cognitive Development (ABCD) study, the largest longitudinal study of brain development in adolescence in the United States, to test if our rodent findings translate to humans (**Figure 3A**).

**Figure 3.**
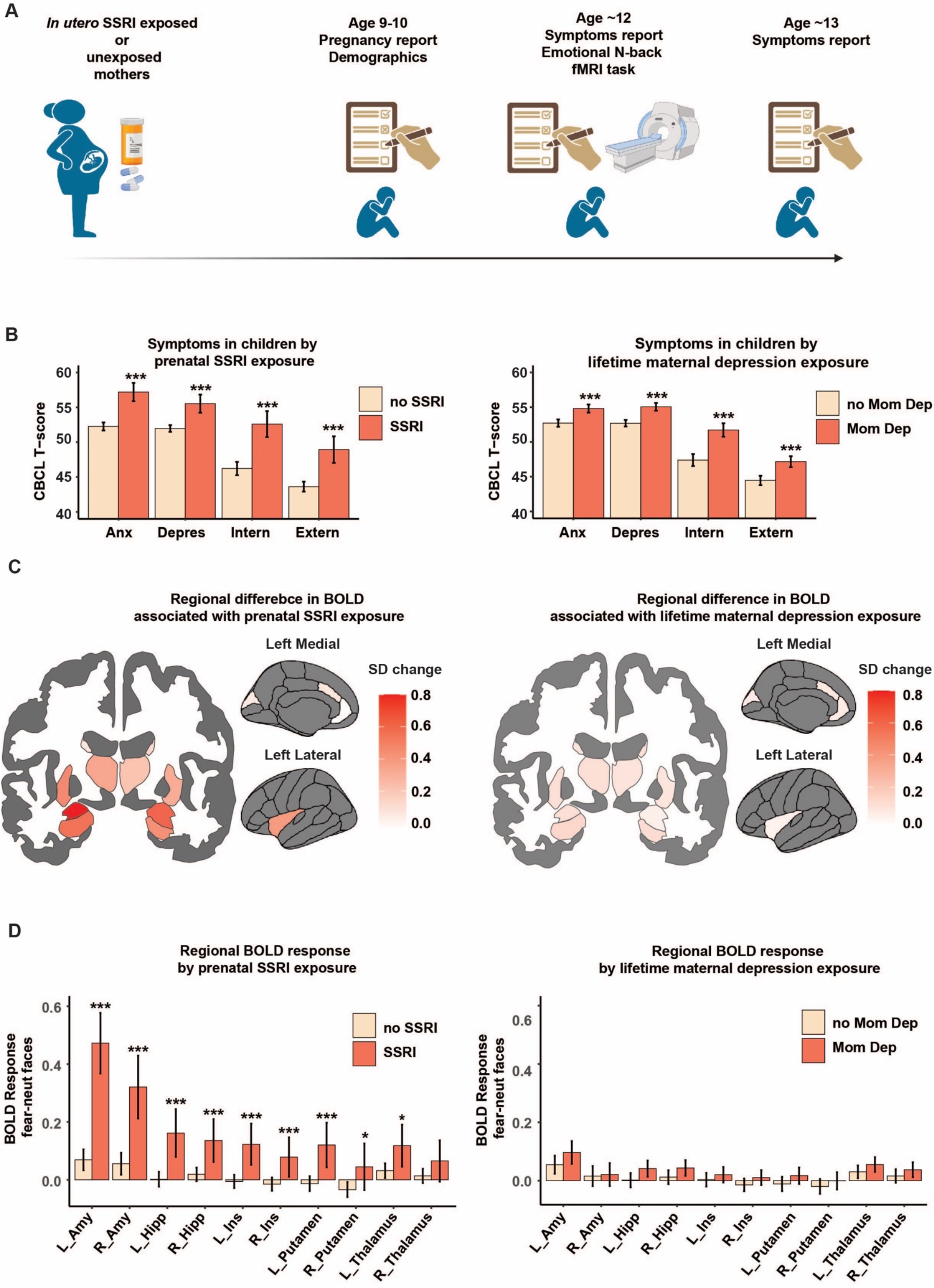
Effects of *in utero* SSRI exposure in human adolescents. **A**) The experimental timeline. At baseline mothers reported on their pregnancy and demographics. Children underwent an MRI scan approximately two years post-baseline, and mothers filled out the Child Behavioral Checklist (CBCL) to assess symptoms at that time and one year post-MRI. **B) Left**. *In utero* SSRI exposure is associated with increased CBCL symptoms, accounting for maternal lifetime depression and maternal anxiety and depressive symptoms. **Right**. Maternal lifetime depression is also associated with increased offspring symptoms. **C) Left**. *In utero* SSRI exposure is associated with increased amygdala, hippocampus, insula, putamen and thalamus activation to fearful-neutral faces, depicted as change in standard deviation of the BOLD response accounting for maternal lifetime depression and maternal anxiety and depressive symptoms. **Right**. Exposure to maternal lifetime depression does not result in significant differences in BOLD response to fearful-neutral faces. **D)** Same analysis as **C**, depicted as predicted means by group, error bars signify prediction intervals. **Left**. *In utero* SSRI exposure is associated with increased BOLD activation to fearful-neutral faces. **Right**. Exposure to maternal lifetime depression does not result in differential BOLD response.

We selected children (**Supplemental Table 3** for demographics) for whom 1) the biological mother was the reporter and had reported on medication use during pregnancy, 2) who had MRI data at age ∼12, 3) concurrent child behavior checklist (CBCL) scores, and 4) complete covariate data (see **Methods**), resulting in N=95 SSRI^+^ children exposed to SSRIs *in utero* and N=3813 SSRI^-^ unexposed children for analyses without MRI data, and in N=68 SSRI^+^ children exposed to SSRIs *in utero* and N=2928 SSRI^-^ unexposed children for analyses containing MRI data. All analyses used propensity score weighting as well as covariates to mitigate potential differences between children of mothers who are treated with SSRIs and those who are not (see **Methods**). We first found that *in utero* SSRI exposure was associated with increased anxiety (b = 0.81, p _FDR_=3.8×10^−19^; betas denote the predicted amount of standard deviations the outcome changes by with SSRI exposure), depressive (b = 0.42, p _FDR_=8.0×10^−7^), internalizing (b = 0.46,p _FDR_=5.6×10^−9^), and externalizing (b = 0.22,p _FDR_=0.006) symptomatology at age 11-13 (**Figure 3B**). As expected from extensive literature showing that maternal depression increases risk for psychiatric disorders in the offspring^41,42^, maternal lifetime depression as a covariate in the same model was also significant, but with smaller effect sizes as indicated by the betas, for anxiety (b = 0.14, p _FDR_=0.003), depressive (b = 0.15, p _FDR_=8.6×10^−4^), internalizing (b = 0.19, p _FDR_=5.4×10^−6^) and externalizing (b = 0.10, p _FDR_=0.034) symptoms. A sensitivity analysis using a model with maternal lifetime depression as the predictor and without prenatal SSRI use or maternal anxiety and depressive symptoms at time of child assessment as covariates also resulted in significant associations with child anxiety (b = 0.35, p _FDR_=1.3×10^−16^), depressive (b = 0.40, p _FDR_=9.1×10^−23^), internalizing (b = 0.42, p _FDR_=4.5×10^−30^) and externalizing (b = 0.27, p _FDR_=5.8×10^−13^) symptoms (**Figure 3B)**.

To test brain activity associated with the innate fear response, we compared BOLD response to fearful versus neutral faces in SSRI^+^ versus SSRI^-^ children. We *a priori* selected seven regions of interest that were significant in the mice (amygdala, rostral and caudal ACC, cuneus, thalamus, caudate and putamen) and two regions that were affected in previous rodent or infant studies (hippocampus and insula). Similar to our findings in mice, SSRI^+^ children had increased response to fearful-neutral faces in the bilateral amygdala (left: b = 0.77, p _FDR_= 4.0×10^−14^; right: b = 0.51, p _FDR_= 2.3×10^−6^; **Figure 3C**,**D**), bilateral hippocampus (left: b = 0.46, p _FDR_= 2.3×10^−6^; right: b = 0.34, p _FDR_= 2.7×10^−4^), bilateral insula (left: b = 0.41, p _FDR_= 6.8×10^−5^; right: b = 0.29, p _FDR_= 0.004), bilateral putamen (left: b = 0.37, p _FDR_= 1.7 ×10^−4^, right: b = 0.24, p _FDR_= 0.029) and left thalamus (b = 0.26, p _FDR_= 0.016). These findings, except for left thalamus and right putamen, remained significant when accounting for concurrent child CBCL scores (**Supplemental Table 4**).

Maternal lifetime depression was controlled for as a covariate in these models and was significantly associated with bilateral cuneus and thalamus, however after FDR correction for multiple comparisons maternal lifetime depression did not remain significantly associated with BOLD response in any region. In a model without covariates for prenatal SSRI exposure and maternal anxiety and depressive symptoms at time of MRI, maternal lifetime depression was not significantly associated with BOLD response to fearful-neutral faces in any region after FDR correction (**Figure 3C**,**D** and **Supplemental Table 5**). To further isolate SSRI effects from maternal depression effects, we tested whether prenatal SSRI use was also associated with differential BOLD response <u>within</u> the subset of children of mothers who reported having a lifetime history of depression (N=44 SSRI^+^ children of mothers with lifetime depression and N=656 SSRI^-^ children of mothers with lifetime depression). In children of mothers with lifetime depression, prenatal SSRI use remained associated with amygdala response (left: b = 0.77, p _FDR_= 1.3×10^−5^; right: b = 0.35, p = 0.03, p _FDR_= 0.16) and left hippocampal response (left: b = 0.34, p _FDR_= 0.038). A last sensitivity analysis comparing three groups: 1) children exposed to SSRI in utero, 2) children exposed to maternal depression who did not take SSRIs during pregnancy, 3) non-exposed children also shows that only the SSRI-exposed group had increased BOLD response (**Supplemental Table 6**), again showing the specificity of the SSRI exposure effects.

Lastly, controlling for prenatal SSRI use and maternal lifetime depression as well as the other covariates, bilateral amygdala response was associated with child anxiety (left: b = 0.12, p _FDR_= 5.4×10^−8^, right: b = 0.07, p _FDR_= 9.9×10^−4^), depressive (left: b = 0.12, p _FDR_= 3.8×10^−9^, right: b = 0.12, p _FDR_= 7.9×10^−9^) and internalizing (left: b = 0.07, p _FDR_=1.7×10^−5^, right: b = 0.09, p _FDR_= 8.0×10^−8^), but not externalizing (left: b = 0.01, p _FDR_= 0.63, right: b = -0.01, p _FDR_= 0.54) symptomatology at time of MRI, as well as with future depressive symptoms one year post MRI (N=62 SSRI^+^,N= 2608 SSRI^-^ with follow-up data; left: b = 0.06, p _FDR_= 0.007, right: b = 0.08, p _FDR_= 4.5×10^−5^). Furthermore, left (but not right) amygdala response to fearful-neutral faces formally mediated the response between SSRI exposure and anxiety symptoms at time of MRI (indirect effect: 0.09, p<0.0001; total effect 0.96, p<0.0001, proportion mediated: 8.7% p<0.0001).

These findings show that similar to the rodent findings, *in utero* SSRI exposure is associated with increased symptomatology, increased fear circuit-related brain activation to innately fearful stimuli and that the amygdala predicts current as well as future depressive symptomatology, suggesting that the amygdala mediates effects by which prenatal SSRI exposure leads to increased risk for psychopathology in the offspring.

### Interpretation

The present findings support the hypothesis that perturbed 5-HT signaling during early development leads to structural and functional changes in both emotional and cognitive domains^12–14,20,33,43,44^ across species and increases risk of psychiatric disorders later in life. PND2-11 SSRI exposure in mice results in reduced motivation to rewarding cues, increased anxiety in elevated plus maze and open field tests, impairments in fear extinction, and depressive-like symptoms in adulthood^20,33,43,45,46^. These wide-ranging behavioral phenotypes in mice align with the increased anxiety, depression, internalizing as well as externalizing symptoms following *in utero* SSRI exposure we observed in human adolescents.

Early in development, the 5-HT transporter is expressed both in serotonergic and non-serotonergic neurons across the brain, while postnatal 5-HT transporter expression is mainly limited to serotonergic neurons as shown in rodents, primates and humans^47–49^. Consequently, SSRIs exert distinct effects during early brain maturation compared to adulthood. The brain structures in the innate fear circuit affected by perinatal SSRIs across species process negative cues and coordinate appropriate threat responses^50–54^. Amygdala and thalamus hyperactivity is likely involved in the behavioral changes (e.g., excessive freezing or internalizing symptoms), because these structures display similar hyperactivation in anxious and depressed individuals even in the absence of awareness^51,55,56^. Moreover, amygdala hyperactivation correlates with current and future symptomatology in humans. Hyperactivity in amygdala and insula could be a consequence of early structural changes in amygdala and insula volume and connectivity observed in human neonates exposed to *in utero* SSRIs^27^. Mice exposed to early life SSRIs exhibit reduced innervation of medial prefrontal cortex (mPFC) by 5-HT fibers^33^ possibly resulting in reduced mPFC top down inhibition of the amygdala underlying exaggerated freezing responses in fear conditioning^57–59^. Whether *in utero* SSRI exposure in humans affects fear circuitry function and psychopathology by decreasing top-down control still remains unknown^60^. Whatever the mechanism, hyperactivity in the insula-amygdala circuit is associated with (increased risk for ^61^) anxiety disorders^62^ which comports with the increased internalizing symptoms observed in SSRI-exposed children in our study. Importantly, amygdala reactivity to faces has been associated with future response to antidepressant medication^63^. Disrupted functioning of the putamen may impair goal-directed behaviors that are important in conflict situations associated with anxiety and depression^64,65^.

Although rodent preclinical models often fail to offer mechanistic insights when crossing phylogeny to humans, our findings indicate that similar developmental mechanisms may be conserved across species. Our experimental mouse model can control for confounding factors, such as maternal depression or treatment, (that typically cloud the interpretation of human data) and allows experimental designs that can establish causality. Subsequent translation to humans shows that our findings are clinically relevant and conserved across species.

Prenatal anxiety and depression adversely affect both mother and child and motivate the public health efforts to identify and treat these disorders. SSRIs are a common therapeutic strategy in perinatal maternal emotional disorders, however the present cross-species data and those from prior studies^4,27,29,66^ indicate that there is a need for studies to develop potentially effective treatments that do not adversely impact the developing fetus in later life. Lastly, our and previous data^22^ indicate symptomatology might begin in early adolescence and suggest that SSRI-exposed children should be screened around this age to direct therapeutic interventions that can prevent the onset or worsening of the behavioral sequelae to those children most at risk.

## Data Availability

ABCD study data is available for download through nda.nih.gov

https://nda.nih.gov/

## Acknowledgements

We would like to thank members of the Ansorge’s lab for methodological and conceptual discussions

## Author Contributions

GZ, MSA, MCC, MvD, AT, JAG, CF, and MMW conceived and supervised all aspects of the study. GZ and MCC performed behavioral and imaging experiments in mice. ASM assisted with mouse behavior. MvD performed behavioral and imaging assessment of human ABCD data. GZ and MvD wrote the manuscript. GGS, ALK, and NP assisted with Matlab script for Deeplabcut analysis of behavior. ALR and ALK assisted with data curation and analysis. CF, MCC, and PK performed and analyzed brain imaging experiments in mice. CLC and PDG contributed conceptually to the human study design. GZ and MvD contributed equally and have the right to list their name first in their CV. All authors contributed to the article and approved the submitted version.

## Data and materials availability

ABCD study data is available for download through nda.nih.gov.

## Supplementary Materials

Materials and Methods

Supplementary Results

Supplementary Figure S1

Tables S1 to S11

References (*67*–*77*)

## Supplementary material

## Materials and Methods

### Mouse study

#### Ethical permission guidelines and subjects

Mice (129SvEv/Tac) were bred at Columbia Psychiatry, New York State Psychiatric Institute. All methods were approved by the Animal Care and Use Committee of the New York State Psychiatric Institute and in accordance with the NIH Guide for the Care and Use of Laboratory Animals according to protocol number 1562. Mice used for experiments were born from litters containing 2–10 pups. Mice were separated by sex and weaned into groups of five mice per cage at P26. Animals were maintained on a 12/12 hr light/dark cycle (lights on at 7:00 a.m.) and provided with food and water *ad libitum*. During the perinatal period, postnatal day 2 to 11 (PND 2-11) mice received daily intraperitoneal injections with either vehicle Saline (Veh; 0.9% NaCl, 5 ml/kg) or fluoxetine (PNFLX; 10 mg/kg in VEH, 5 ml/kg). Multiple cohorts of mice were created for the various experiments. For innate fear behavioral assessment, we used female mice from at least 4 litters and 7 individuals in each treatment group. For adolescent fear behavior we used Pet1Cre mice on a 129SvEv/Tac background. For fMRI experiments we used Male Saline n=8, Male PNFLX n=13, Female Sal n =10, Female PNFLX n=13.

#### Innate fear behavior tests

Adult mice were handled for a week prior to testing to get acquainted with the experimenter. Innate fear behavior was tested on two consecutive days: habituation and test.

##### Fear apparatus in adulthood

On the day of testing animals were brought individually into the behavior room in a transport cage. The testing apparatus consisted of a 3-compartment chamber opened on the top with air flowing from one side and an air suction system positioned on the opposite side, creating a flow left to right. The apparatus floor was covered in corn cob bedding. The size of the apparatus was in cm 64 (L) x 43.8 (W) x 30.5 (H) and 4 separators placed at equal distance of size in cm 11 (W) x 30.5 (H) that defined 3 zones: safe (farthest from the predator/neutral odor), neutral (middle and adjacent to both predator and safe zones), and predator (closest to the predator/neutral odor). The animals were tested individually for 10 minutes during habituation and allowed to explore the 3-chamber maze starting from the neutral zone where the animal was originally placed in the absence of the predator odor. On the following day the animals were tested for 10 minutes in presence of a predator or neutral odor placed at the right respective opposite corners of the predator odor zone. The predator odor consisted of mountain lion urine (LLC 92012) that was dispensed through soaked cue tips inserted in a plastic holder taped at the two right respective opposite corners of the predator zone. The neutral odor consisted of a commercial banana scent (McCormick). The animal did not interact with the odor directly and we used 100 µl for each cue tip that was changed between animals. We observed behavioral effects that indicated rapid diffusion of the odor in the entire chamber, so we also examined overall behavior, irrespective of compartment position (data not shown).

##### Fear testing in adolescence

On the day of testing animals were brought individually into the behavior room in a transport cage. The testing apparatus consisted of an air-tight clear plastic chamber that has a clear lid (12 1/2 ‘‘L x 8 1/2 ‘‘W x 8 1/2 ‘‘H). A predator-like odor, 2-methyl-2-thiazoline (2MT), was delivered in an odor-attenuated air flow-controlled chamber. Briefly, 2MT air or odor-free air was administered via a stopcock valve connected two Erlenmeyer flasks via different tubing lines. The odor was introduced in the chamber by placing a piece of filter paper scented with 2MT (3µl) at the top of the flask connected with the odor delivery line. Continuous airflow was maintained through the airtight chamber at 4 psi via calibration columns. The fear odor experiment was performed under a laminar flow hood and airflow is exhausted from the building. The 2MT has consistent formulation and high potency, and this odor has been previously shown to induce measurable levels of fear-like behaviors in mice amounting to 80-90% ^1–3^. The testing started with 2 minutes of air followed by 3 minutes of 2MT, and 10 more minutes of air (**Figure 1 Supplementary**).

##### Behavioral scoring and video analysis

A top view ANYmaze camera (Digital USB 2.0 CMOS Camera, DMK 22AUC03) was connected to the ANYmaze software and the animal’s body was tracked throughout habituation and test tasks in both exposures. All videos in adulthood and adolescence were analyzed offline using ANYmaze software. The parameters included in the ANYmaze analysis were total time immobile (3 seconds immobility threshold), total distance traveled, latency to immobility, time in each zone, number of entries in each zone, average speed in each zone. Additional parameters are reported in Figure 1 supplementary: latency to first enter safe and predator zones, total immobile episodes, total mobile time in each zone, and total immobile time in each zone.

##### DeepLabCut pose estimation

We conducted a more granular analysis of the adult innate fear behavioral response in our animal model using DeepLabCut ^4^. This software enables estimation of poses by tracking the animal body parts with high accuracy. Data was recorded at 30 Hz by one camera: the 640 × 480 pixels images were acquired with a Digital USB 2.0 CMOS Camera, DMK 22AUC03. Since our behavior was uniform overtime, we extracted 20 labels using K-Means clustering and two independent human annotators blinded to the treatment were trained to localize the following body parts: nose, left and right ear, left and right hip, tailbase, and tailend. The tracking consisted of randomly extracted frames for a total of 280 frames from 14 mice scored separately for habituation and predator odor. We then created a training dataset using a resnet_50 network and default augmentation method. To train the network we used a shuffle=1, training index=0, and a 150k maximum iterations, the point at which the training reached a plateau. We used custom MATLAB scripts to analyze the tracked videos produced from DeepLabCut (R2019a.Ink, version 9.6.0.1072779). We plotted the likelihood data simultaneously with the tracked videos to visually determine a confidence threshold (0.95) of the labeled points. Outliers were removed if they were more than three scaled median absolute deviations (MAD) away from the median. Distances between body parts were calculated using the respective x and y coordinates for each analyzed video frame. Body perimeter was calculated using Heron’s formula to calculate triangle perimeter and by summing the perimeter of the triangles formed by the following body parts distances: nose-left ear-right ear, left ear-right ear-left hip, right ear-left ear-right hip, tail base-right hip-left hip. Data is presented in pixels.

#### Functional magnetic resonance imaging (fMRI) during predator odor exposure

##### Awake Mouse Imaging System

Presented in **Figure 1A** are the different components of the mouse imaging system showing a radiofrequency coil and MR compatible restraining system for imaging awake mice (Ekam Imaging, Boston MA USA). The quadrature transmit/receive volume coil (ID 38 mm) provides excellent anatomical resolution, signal-noise-ratio (SNR) for voxel-based fMRI. The unique design of the holder essentially stabilizes the head in a cushion, minimizing any discomfort normally caused by ear bars and pressure points used to immobilize the head for awake animal imaging. Odorants are delivered through PE tubing connected to the tubular bite bar used to secure the front incisors of the mouse in the head holder. A movie showing the set-up of a mouse for awake imaging is available at http://www.youtube.com/watch?v=W5Jup13isqw.

##### Acclimation

A week prior to the first imaging session, all mice were acclimated to the imaging system before scanning. Mice were secured into their holding system while anesthetized with 1-2% isoflurane. Following cessation of isoflurane, fully conscious mice were put into a ‘mock scanner’ (a black box with a tape recording of MRI pulses) for 30 minutes for four consecutive days. Acclimation in awake animal imaging significantly reduces physiological effects of the autonomic nervous system including heart rate, respiration, corticosteroid levels, and motor movements helping to improve contrast-to-noise and image quality ^5,6^.

##### Imaging Acquisition and Pulse Sequence

Experiments were conducted using a Bruker Biospec 7.0T/20-cm USR horizontal magnet (Bruker, Billerica, Massachusetts) and a 20-G/cm magnetic field gradient insert (ID = 12 cm) capable of a 120-µs rise time (Bruker). At the beginning of each imaging session, a high-resolution anatomical data set was collected using the RARE pulse sequence (20 slice; 0.75 mm; FOV 2.5 cm; data matrix 256 × 256; TR 2.1 sec; TE 12.4 msec; Effect TE 48 msec, NEX 6; 6.5 min acquisition time). Functional images were acquired using a multi-slice HASTE pulse sequence (<u>H</u>alf Fourier <u>A</u>cquisition <u>S</u>ingle Shot <u>T</u>urbo Spin <u>E</u>cho). With this sequence it is possible to collect twenty, 0.75 mm thick, axial slices in less than six seconds. With a FOV of 2.5 cm and a data matrix of 96 × 96, the in-plane pixel functional resolution for these studies was 260 µm^2^.

##### Provocation Paradigm - Odor Stimulant

Awake mice were imaged for changes in BOLD signal intensity in response to the odor of mountain lion urine (PredatorPee Liquid, www.predatorpee.com). The control scent was bedding from the home cage.

##### Data Analysis

Images were aligned and registered to a 3D mouse brain atlas, which is segmented and labeled with 134 discrete anatomical regions (Ekam Solutions, Boston MA). The alignment process was facilitated by an interactive graphic user interface. The registration process involved translation, rotation and scaling independently and in all three dimensions. Matrices that transformed each subject’s anatomy were used to embed each slice within the atlas. All pixel locations of anatomy that were transformed were tagged with major and minor regions in the atlas. This combination created a fully segmented representation of each subject within the atlas. The inverse transformation matrix [Ti]-1for each subject (i) was also calculated.

In voxel-based analysis, the BOLD % change of each independent voxel was averaged for all subjects. Statistical t tests were performed on each voxel (ca. 15,000 in number) of each subject within their original coordinate system with a baseline threshold of 2% BOLD change to account for normal fluctuation of BOLD signal in the awake rodent brain ^7^. As a result of the multiple t test analyses performed, a false-positive detection controlling mechanism was introduced ^8^. This subsequent filter guaranteed that, on average, the false-positive detection rate was below our cutoff of 0.05. The t test statistics used a 95% confidence level, two-tailed distributions, and heteroscedastic variance assumptions.

A composite image of the whole brain representing the average of all subjects was constructed for each group for ROI analyses, allowing us to look at each ROI separately to determine the BOLD change and the number of activated voxels in each ROI. Statistical comparisons of different image acquisitions are compared to baseline. A non-parametric Kruskal-Wallis test statistic was used to compare the average signal intensity in each of ca 15,000 voxel for their first 3.0 minutes baseline (acquisitions 1-30) to minutes 7-12 (acquisitions 70-120).

### Statistical analysis

Data was analyzed using GraphPad Prism (version 8.4.3). Repeated measures two-way ANOVA and three-way ANOVA were used for analysis of variance of Treatment (vehicle vs perinatal fluoxetine), Exposure (habituation vs test), and Time variables. When interactions of variables were significant a multiple comparison post hoc test was conducted, and significance was reported in the graph. Significant main effects of individual variables were also reported in the graph. Outliers were identified using the ROUT method using a Q coefficient of 1% and the pairwise comparison was removed from the analysis. Statistical data output for each test performed are summarized in Table 1, Table 2, and Table 1 and 2 supplementary. All data are presented as the mean ± SEM. Significance was \**p<0*.*05*; \*\**p<0*.*01*; \*\*\**p<0*.*001*.

### Human Study

#### ABCD study data

For the human experiments we did secondary analyses of Adolescent Brain Cognitive Development^SM^ Study (ABCD Study®) data, a large longitudinal nationwide study of children recruited from primary and public-school systems at 21 sites nationwide. Since phenotypes may only appear in adolescence ^9^, we used the latest MRI data available, the two-year follow up data, when the children were 10.6-13.8 years old. The analysis sample was from the NDA’s two year follow-up release (downloaded through October 2022) and had the following additional inclusion criteria: biological mother reporting, data available about prenatal medication use and other prenatal and birth history as well as full data on other covariates (see below) and two year follow-up data available from the parent-report child behavior checklist (CBCL), which resulted in N=95 SSRI^+^ children exposed to SSRIs *in utero* and N=3813 SSRI^-^ unexposed children. For analyses including an association with BOLD response we also required complete data on the N-BACK fMRI task including behavior and that MRI scans passed quality control measures as advised by the ABCD study, resulting in N=68 SSRI^+^ children exposed to SSRIs *in utero* and N=2928 SSRI^-^ unexposed children. ABCD study procedures were approved by the Institutional Review Board at the University of California at San Diego, San Diego, CA. Parents provided written informed consent; children provided verbal assent. The local New York State Psychiatric Institute IRB approved secondary analyses of ABCD data

#### Procedures and Variables of Interest

Biological mothers completed reports on demographics, prenatal history, CBCL about their child, adult self report (the equivalent to CBCL for adults, about themselves), and their own psychiatric history through the family history assessment (see **Supplemental Table 11** for detailed variables).

#### MRI

As reported in detail elsewhere^10^ children performed the emotional N-back task in the MRI scanner. Briefly, children viewed images of positive (happy), negative (fearful) or neutral emotional faces and indicated if the current image was the same as the previous image (0-back) or as the image they saw two images earlier (2-back). We used preprocessed data made available by the ABCD study comparing BOLD response to negative compared to neutral images in a-priori selected regions of interest amygdala, hippocampus, putamen, insula, rostral and caudal ACC, cuneus and thalamus. These regions were selected to translate the mouse findings (Figure 2) and/or because in earlier work with neonatal infants ^11^ we had found that the amygdala and the insula were affected.

#### Measures

##### Predictor: prenatal SSRI use

Data regarding SSRI use once mothers knew that they were pregnant were used to make a composite SSRI use variable. Children were grouped as exposed to SSRI if mothers reported having used while they knew they were pregnant the following: fluoxetine, fluvoxamine, sertraline, citalopram, escitalopram, and/or paroxetine, but did not also use other types of antidepressants such as bupropion, duloxetine, MAO-inhibitors, tricyclic antidepressants, anti-anxiety medication, opioids, antipsychotic medication, anti-convulsant medication, stimulants or sleep aids.

Because the SSRI exposure had the largest effect size on the amygdala response, we then selected bilateral amygdala BOLD response to fearful versus neutral faces as a predictor for symptomatology at time of MRI and at the 1 year time point after MRI (N= 2670 had this follow-up data available).

##### Potential Confounders

All our analyses controlled for birth weight, child age at time of MRI, sex assigned at birth, puberty score at time of MRI, maternal lifetime depression, maternal depression and anxiety symptomatology at time of child MRI (ASR), maternal age at birth, maternal race/ethnicity, area deprivation index, maternal education, combined household income, birth complications, doctors visits during pregnancy, pregnancy illnesses, prenatal vitamin use, prematurity, whether the pregnancy was planned, delivery by cesarean, substance use knowing of pregnancy of the following substances: caffeine, tobacco, alcohol, cannabis and cocaine and crack cocaine, oxycodone. In addition, we added in random effects terms for MRI scanner site and family relatedness.

For analyses concerning the emotional N-back fMRI task we additionally controlled for behavior on the task for neutral and fearful images (rate of correct responses and reaction time). We performed sensitivity analyses and added in child CBCL anxiety, depressive, internalizing and externalizing symptoms to ensure that effects were not due to potential differences in symptomatology. To further ensure that the findings were not solely due to differences in maternal depression between groups, we selected the subsample of children whose mothers reported they had ever been depressed and repeated analyses of prenatal SSRI exposure on BOLD response in the subsample. We also performed analyses directly comparing three groups: *in utero* SSRI exposed children, children exposed to maternal depression but not SSRIs and unexposed children.

### Statistical analyses

Analyses were performed using R (version 1.4.1717). For descriptive analyses, chi-square tests were used for categorical variables, and *t*-tests for continuous variables. For regression analysis linear mixed effects models were used using R-package “lme4”. Analyses were adjusted for multiple comparisons (9 brain regions x 2 hemispheres) with FDR corrections. All continuous variables were standardized, binary variables were not. All analyses were weighted by propensity scores to minimize confounding due to differences between mothers who used SSRIs prenatally and mothers who did not (see below). The covariates that Individual BOLD response values were winsorized to 3 standard deviations from the mean to diminish the influence of outliers. Multicollinearity was of no concern with maximum variance inflation factor quantifying any multicollinearity was 3.5 (for behavioral measures on the N-back task), where 1 is the minimum and values of 10 and higher indicate concerning multicollinearity ^12^. Removing the behavioral N-back variables from the models did not change the results. Mediation analyses was performed including covariates and propensity score weighting using R-package “mediation” for causal mediation analysis.

#### Propensity score weighting

We used propensity score weighting to generate balancing weights for causal effect estimation. We used R-package Weightit to balance prenatal covariates between SSRI exposure and control groups using an ATT (average treatment effect on the treated) estimand. The list of covariates we balanced were the following: knowing of pregnancy use of: alcohol, tobacco, cannabis, caffeine, cocaine/crack cocaine, heroine/morphine, oxycodone, prenatal vitamins, pregnancy illnesses, whether the pregnancy was planned, maternal lifetime depression, maternal age at pregnancy, number of doctors visits during pregnancy, mother’s race/ethnicity, maternal education. The resulting propensity scores were used to weight participants for all linear mixed models. These variables were also entered into the linear mixed models in a double robust approach^13^.

## Supplementary results

### Translation to human adolescent children from the ABCD study

To test if there were any sex effects we entered an interaction term between prenatal SSRI use and child sex into the models and found a significant effect. Stratified analyses showed that BOLD response differences to negative compared to neutral faces were significant in girls but not in boys after FDR correction (**see Supplemental Table 7 and 8**). Similarly, there was a significant interaction between SSRI use in pregnancy and child sex in predicting anxiety, depressive and internalizing symptom scores; stratifying by sex again showed that SSRI use in pregnancy predicted depressive and internalizing symptoms in girls but not in boys (**see Supplemental Tables 9 and 10)**.

## Figure 1 supplementary

**Figure 1_Supplementary.**
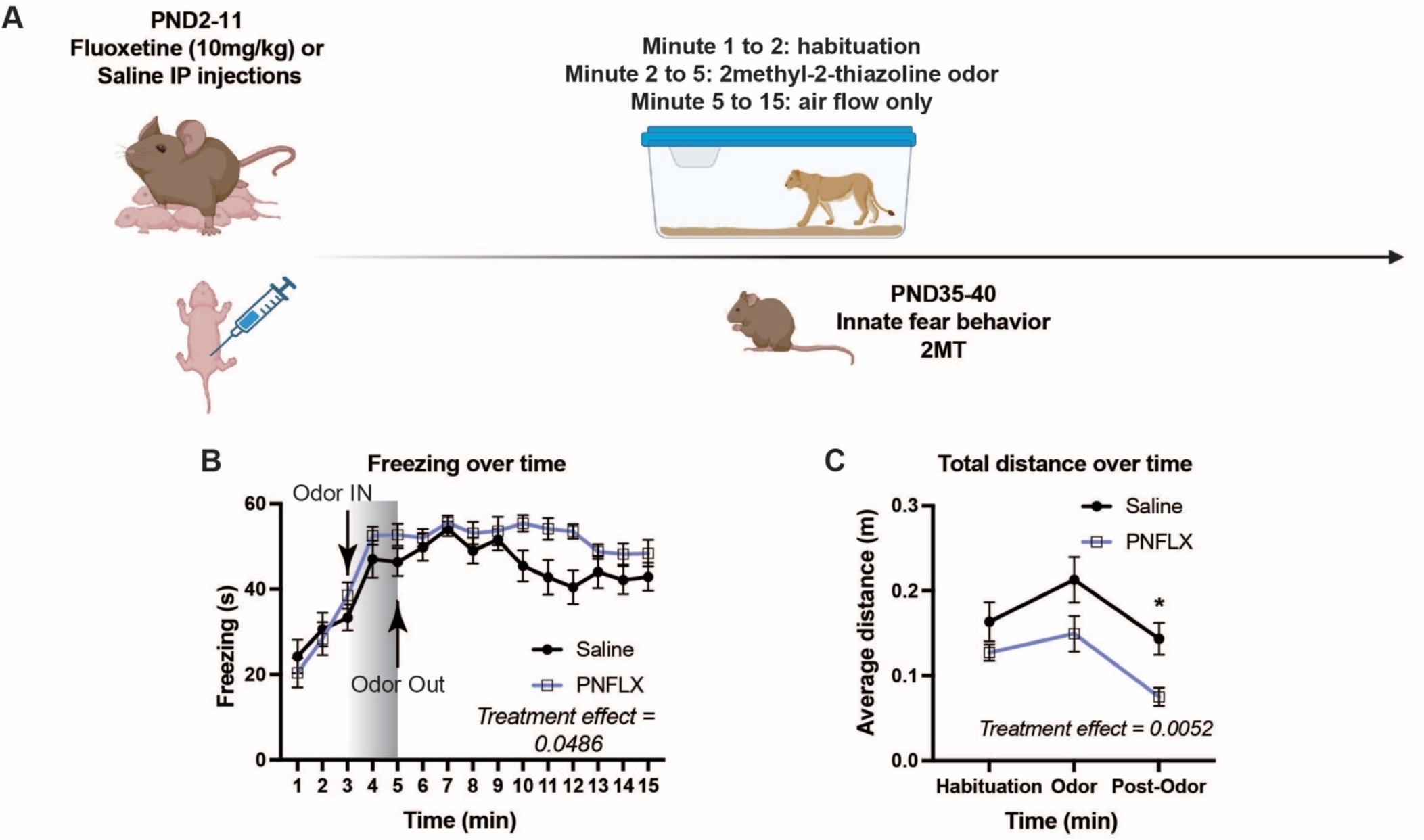
Adolescent exposure to a predator odor increased immobility and crouching in PNFLX treated mice compared to saline mice. **A**)Timeline of the adolescent mouse study. **B**) Time course of immobility over the 15 minutes of 2MT exposure shows that PNFLX mice compared to saline mice are more immobile. **B**) PNFLX mice covered a shorter average distance than saline mice over time. Saline N=9, PNFLX N=10. \**p<0*.*05*; \*\**p<0*.*01*; \*\*\**p<0*.*001*

## Supplemental Tables

**Supplemental Table 1.** Statistical data output from Figure 1.

|  | ANOVA/REML table | SS | DF | MS | F (DFn, DFd) | P value | Outliers |
| --- | --- | --- | --- | --- | --- | --- | --- |
| Total time freezing | **Time** | **1923** | **4** | **480.8** | **F (4, 48) = 4.851** | **P=0.0023** | 0 |
|  | **Test** | **1587** | **1** | **1587** | **F (1, 12) = 3.196** | **P=0.0991** |  |
|  | **Treatment** | **1053** | **1** | **1053** | **F (1, 12) = 4.302** | **P=0.0602** |  |
|  | Time x Test | 351.4 | 4 | 87.85 | F (4, 48) = 1.124 | P=0.3564 |  |
|  | **Time x Treatment** | **1844** | **4** | **461** | **F (4, 48) = 4.651** | **P=0.0030** |  |
|  | **Test x Treatment** | **2565** | **1** | **2565** | **F (1, 12) = 5.168** | **P=0.0422** |  |
|  | Time x Test x Treatment | 152.2 | 4 | 38.04 | F (4, 48) = 0.4866 | P=0.7455 |  |
| Latency to freeze | **Test x Treatment** | **2421** | **1** | **2421** | **F (1, 10) = 5.417** | **P=0.0422** | 2 |
|  | **Test** | **19875** | **1** | **19875** | **F (1, 10) = 44.46** | **P<0.0001** |  |
|  | **Treatment** | **2421** | **1** | **2421** | **F (1, 10) = 5.417** | **P=0.0422** |  |
| Total distance traveled | **Time** | **31.34** | **4** | **7.835** | **F (4, 48) = 10.90** | **P<0.0001** | 0 |
|  | Treatment | 0.193 | 1 | 0.193 | F (1, 12) = 0.1091 | P=0.7469 |  |
|  | Test | 33.43 | 1 | 33.43 | F (1, 12) = 8.844 | P=0.0116 |  |
|  | Time x Treatment | 0.8282 | 4 | 0.2071 | F (4, 48) = 0.5482 | P=0.7012 |  |
|  | Time x Test | 2.707 | 4 | 0.6768 | F (4, 48) = 0.9417 | P=0.4480 |  |
|  | Treatment x Test | 1.374 | 1 | 1.374 | F (1, 12) = 0.7765 | P=0.3955 |  |
|  | Time x Treatment x Test | 2.164 | 4 | 0.5411 | F (4, 48) = 1.432 | P=0.2377 |  |
| Correlation freezing vs perimeter | Correlation |  |  |  | F (1, 26) = 6.129 | P=0.0201 | 0 |
|  | R squared |  |  |  |  | 0.1908 |  |
| Total average perimeter | Time |  |  |  | F (4, 51) = 0.7792 | P=0.5439 | 2 |
|  | Treatment |  |  |  | F (1, 51) = 0.9200 | P=0.342 |  |
|  | **Time x Treatment** |  |  |  | F (4, 51) = 2.858 | **P=0.0326** |  |
| Total time freezing | **Time** | **1696** | **4** | **424** | **F (4, 48) = 12.51** | **P<0.0001** | 0 |
|  | Treatment | 51.61 | 1 | 51.61 | F (1, 12) = 0.4611 | P=0.5100 |  |
|  | **Test** | **1095** | **1** | **1095** | **F (1, 12) = 28.17** | **P=0.0002** |  |
|  | Time x Treatment | 56.78 | 4 | 14.2 | F (4, 48) = 0.4189 | P=0.7942 |  |
|  | Time x Test | 128.4 | 4 | 32.09 | F (4, 48) = 0.8717 | P=0.4878 |  |
|  | Treatment x Test | 4.393 | 1 | 4.393 | F (1, 12) = 0.1130 | P=0.7426 |  |
|  | Time x Treatment x Test | 242.5 | 4 | 60.62 | F (4, 48) = 1.647 | P=0.1780 |  |
| Latency to freeze | Exposure x Treatment | 3924 | 1 | 3924 | F (1, 12) = 1.838 | P=0.2002 | 0 |
|  | Test | 2146 | 1 | 2146 | F (1, 12) = 1.005 | P=0.3359 |  |
|  | Treatment | 3924 | 1 | 3924 | F (1, 12) = 1.838 | P=0.2002 |  |
| Total distance traveled | **Time** | **60.64** | **4** | **15.16** | **F (4, 48) = 40.05** | **P<0.0001** | 0 |
|  | Treatment | 2.695 | 1 | 2.695 | F (1, 12) = 0.5583 | P=0.4693 |  |
|  | **Test** | **48.67** | **1** | **48.67** | **F (1, 12) = 10.27** | **P=0.0076** |  |
|  | Time x Treatment | 4.204 | 4 | 1.051 | F (4, 48) = 1.816 | P=0.1411 |  |
|  | Time x Test | 9.337 | 4 | 2.334 | F (4, 48) = 6.166 | P=0.0004 |  |
|  | Treatment x Test | 3.52 | 1 | 3.52 | F (1, 12) = 0.7290 | P=0.4099 |  |
|  | Time x Treatment x Test | 1.722 | 4 | 0.4304 | F (4, 48) = 0.7438 | P=0.5669 |  |

**Supplemental Table 2.** Statistical data output from Figure 1 Supplementary.

| **Figure 1 Supp.** | **ANOVA/REML table** | **SS** | **DF** | **MS** | **F (DFn, DFd)** | **P value** |
| --- | --- | --- | --- | --- | --- | --- |
| **B** | **Time** | **21833** | **14** | **1560** | **F (6.745, 114.7) = 23.16** | **p<0.0001** |
|  | **Treatment** | **1616** | **1** | **1616** | **F (1, 17) = 4.513** | **p=0.0486** |
|  | Time x Treatment | 1381 | 14 | 98.66 | F (14, 238) = 1.465 | p=0.1250 |
| **C** | **Time** | **0.04895** | **2** | **0.02447** | **F (1.883, 32.01) = 8.343** | **P=0.0015** |
|  | **Treatment** | **0.04467** | **1** | **0.04467** | **F (1, 17) = 10.29** | **P=0.0052** |
|  | Time x Treatment | 0.002799 | 2 | 0.001399 | F (2, 34) = 0.4770 | P=0.6247 |

**Supplemental Table 3.**
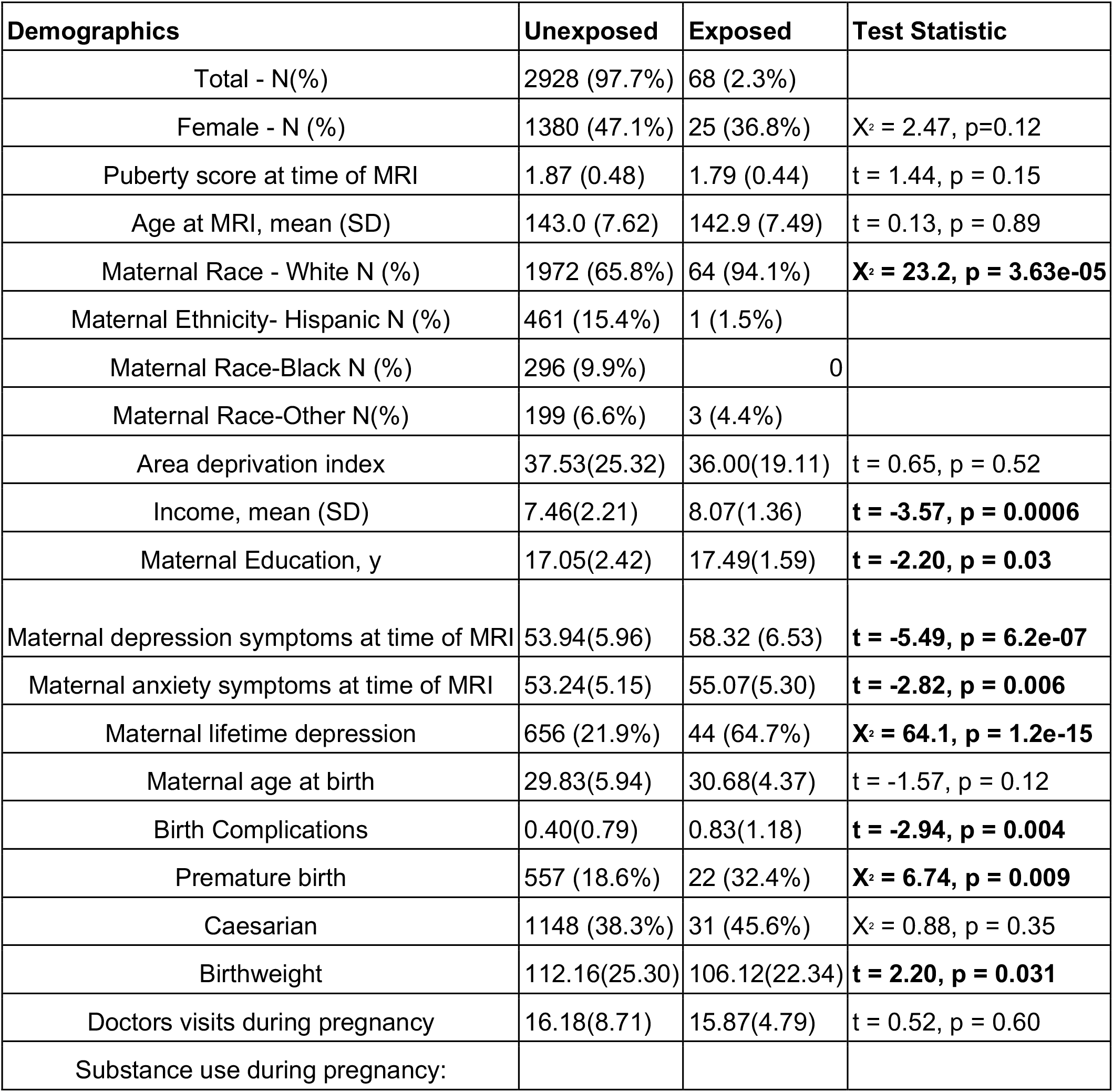
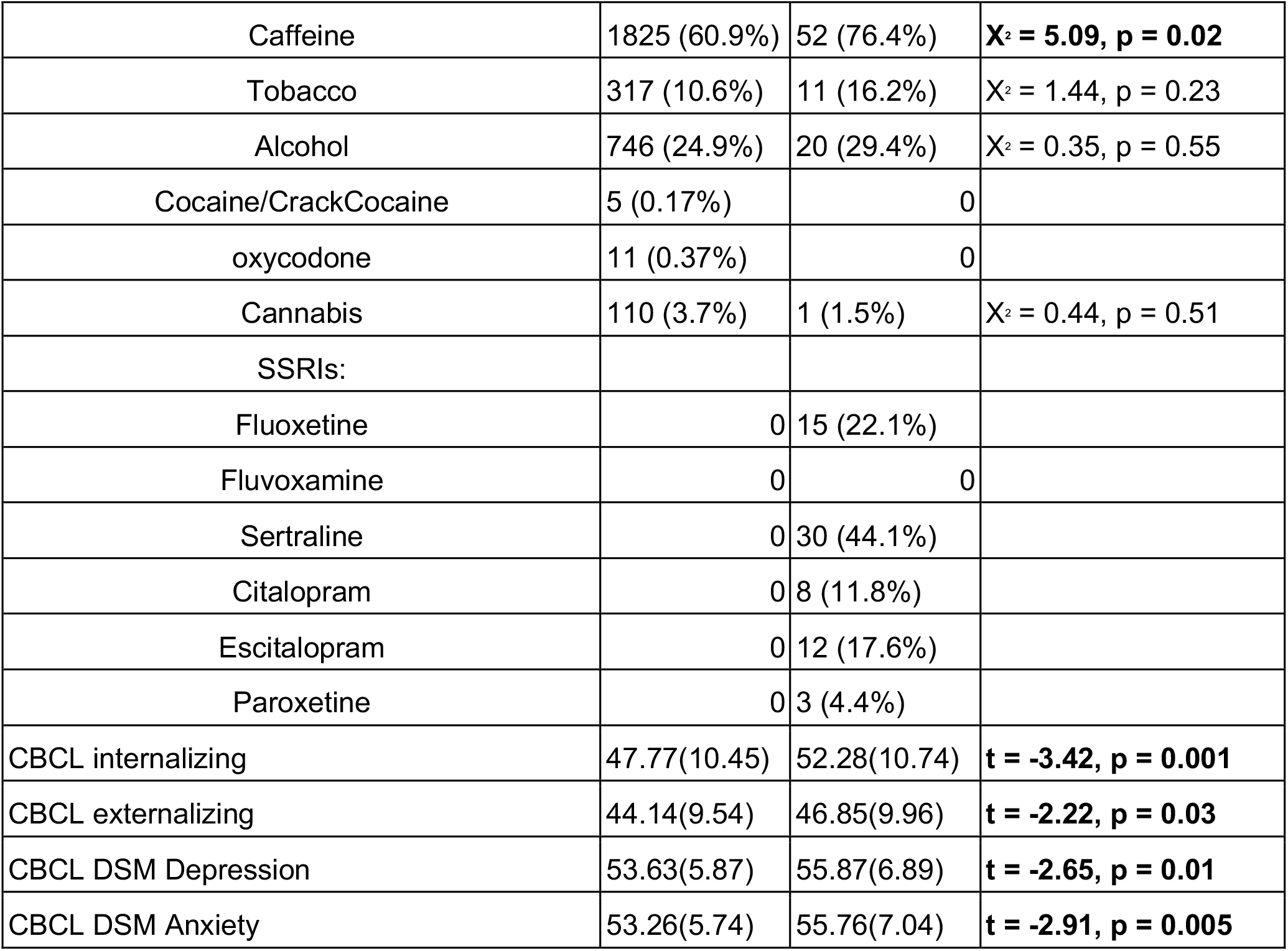
Demographics Table.

**Supplemental Table 4.** Controlling for CBCL scores, in utero SSRI exposure remains associated with increased BOLD response to Negative-Neutral faces compared to unexposed children.

| Brain Region | Beta value | p-value | p-FDR |
| --- | --- | --- | --- |
| Left Amygdala | **0.680608** | 0.000001 | 1.35E-10 |
| Right Amygdala | **0.483497** | 0.000003 | 2.19E-05 |
| Left Hippocampus | **0.426314** | 0.000004 | 2.19E-05 |
| Right Hippocampus | **0.289007** | 0.001176 | 3.63E-03 |
| Left Insula | **0.341187** | 0.000315 | 1.47E-03 |
| Right Insula | **0.240463** | 0.011016 | 2.86E-02 |
| Left Cuneus | 0.027196 | 0.769436 | 8.78E-01 |
| Right Cuneus | 0.019392 | 0.830021 | 8.79E-01 |
| Left Thalamus | 0.204565 | 0.038841 | 8.78E-02 |
| Right Thalamus | 0.117437 | 0.208786 | 3.76E-01 |
| Left Putamen | **0.304776** | 0.001166 | 3.63E-03 |
| Right Putamen | 0.175985 | 0.067091 | 1.35E-01 |
| Left rostral anterior cingulate cortex | -0.079676 | 0.411302 | 6.17E-01 |
| Right rostral anterior cingulate cortex | -0.02673 | 0.780306 | 8.78E-01 |
| Left Caudal ACC | 0.046188 | 0.62392 | 8.60E-01 |
| Right Caudal ACC | 0.089496 | 0.345232 | 5.65E-01 |
| Left Caudate | -0.00741 | 0.934179 | 9.34E-01 |
| Right Caudate | 0.037908 | 0.668925 | 8.60E-01 |

**Supplemental Table 5.** Maternal lifetime depression is not associated with BOLD response to Negative-Neutral faces in children. This model does not control for prenatal SSRI use or maternal symptoms at time of child MRI, and adding those back into the model does not change the results.

| Brain Region | Beta value | p-value | p-fdr |
| --- | --- | --- | --- |
| Left Amygdala | 0.080422 | 0.051238 | 0.1190346 |
| Right Amygdala | 0.011142 | 0.792731 | 0.7927534 |
| Left Hippocampus | 0.112995 | 0.007408 | 0.1190346 |
| Right Hippocampus | 0.091598 | 0.023118 | 0.1190346 |
| Left Insula | 0.054277 | 0.190139 | 0.2190422 |
| Right Insula | 0.078418 | 0.056846 | 0.1190346 |
| Left Cuneus | 0.073827 | 0.064834 | 0.1190346 |
| Right Cuneus | 0.083416 | 0.038816 | 0.1190346 |
| Left Thalamus | 0.076543 | 0.069893 | 0.1190346 |
| Right Thalamus | 0.063446 | 0.116163 | 0.1610481 |
| Left Putamen | 0.080228 | 0.054156 | 0.1190346 |
| Right Putamen | 0.053502 | 0.206736 | 0.2190422 |
| Left rostral anterior cingulate cortex | 0.07485 | 0.072612 | 0.1190346 |
| Right rostral anterior cingulate cortex | 0.053917 | 0.19545 | 0.2190422 |
| Left Caudal ACC | 0.087263 | 0.035586 | 0.1190346 |
| Right Caudal ACC | 0.082964 | 0.047661 | 0.1190346 |
| Left Caudate | 0.052752 | 0.202768 | 0.2190422 |
| Right Caudate | 0.069327 | 0.08982 | 0.1349364 |

**Supplemental Table 6.**
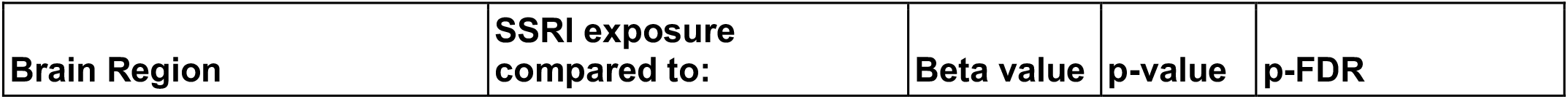
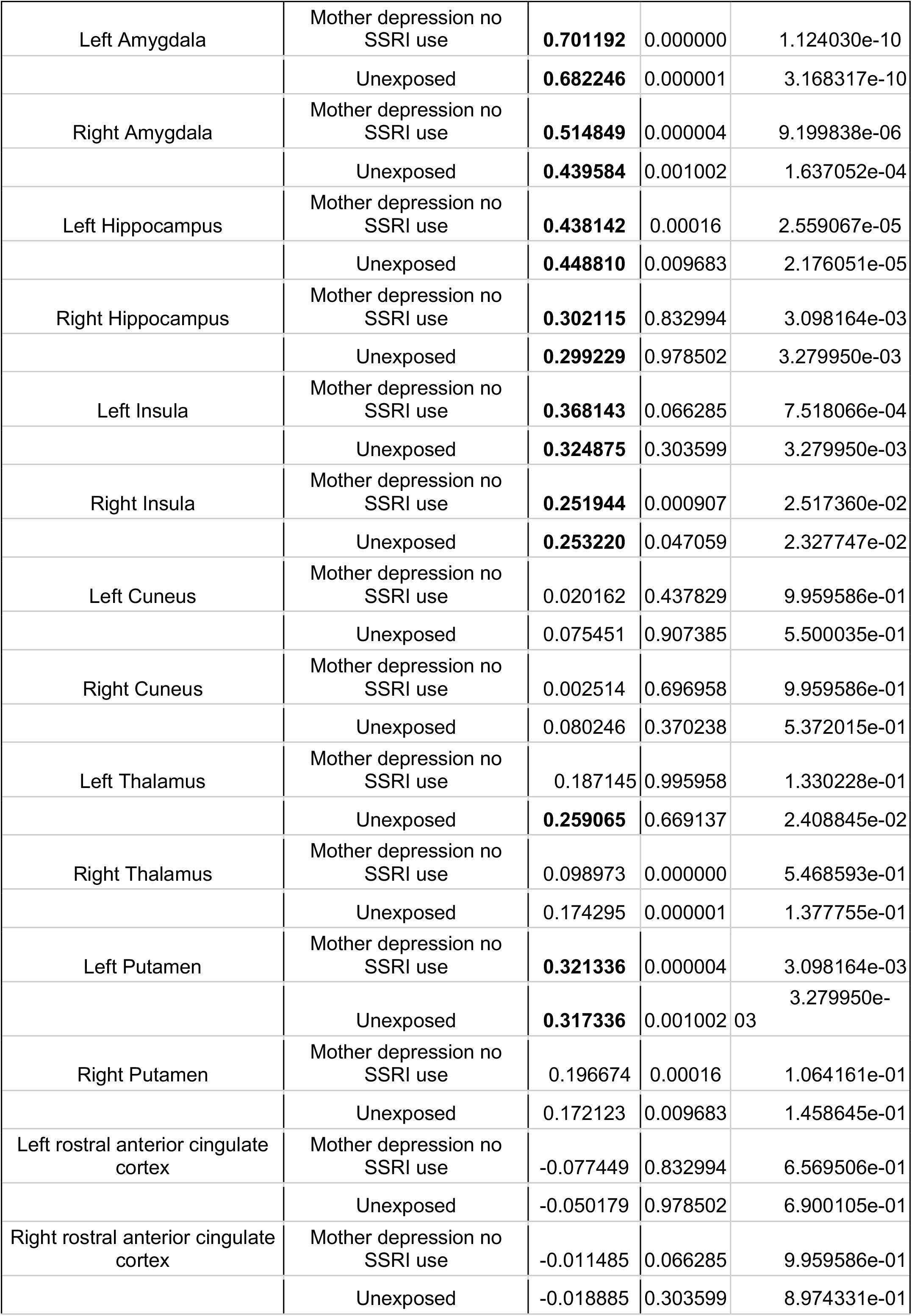
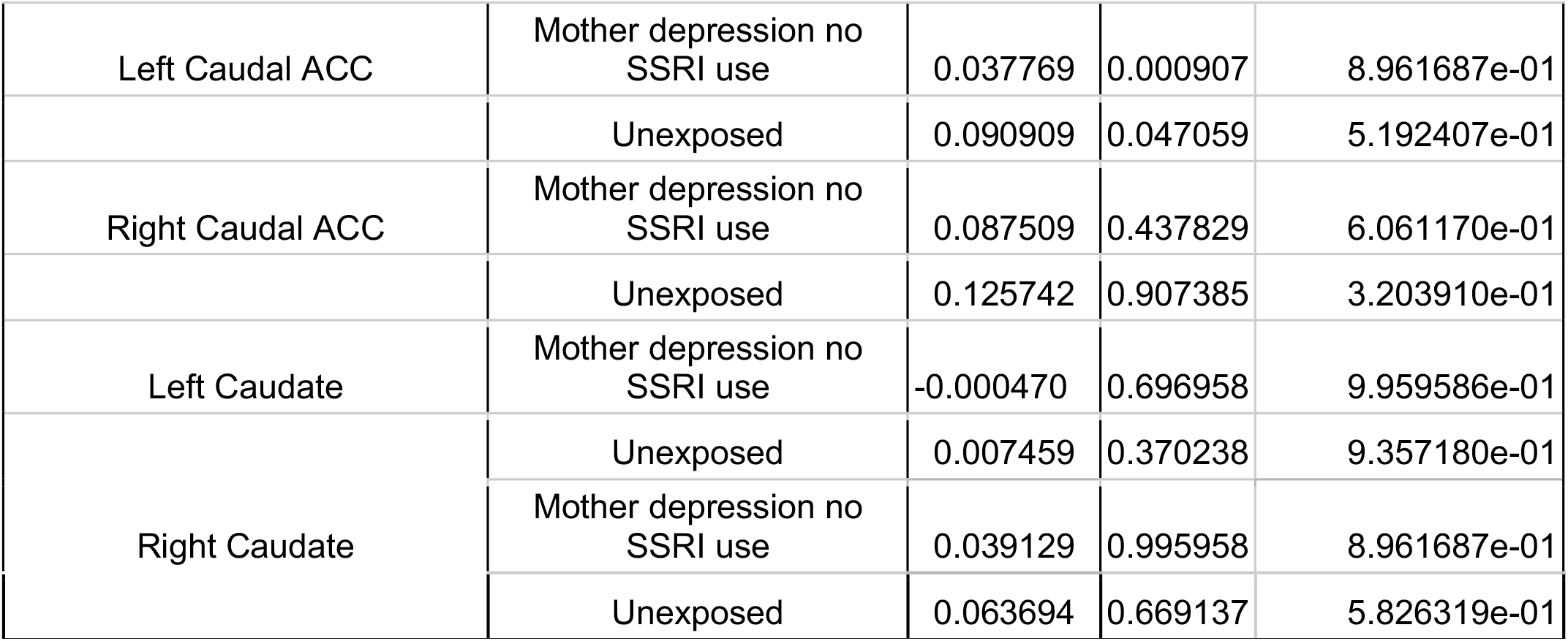
In utero SSRI exposure is associated with increased BOLD response to Negative-Neutral faces in children compared to children of mothers with lifetime depression but no prenatal SSRI use or unexposed (no maternal depression nor in utero SSRI exposure) children.

**Supplemental Table 7.** SSRI exposure is associated with symptoms in girls.

| Symptoms-Girls | Beta value | p-value | p-FDR |
| --- | --- | --- | --- |
| Anxiety | **1.116310** | p = 0.000000 | 6.69E-15 |
| Depressive | **0.479643** | p = 0.000071 | 1.10E-04 |
| Internalizing | **0.566938** | p = 0.000003 | 6.77E-06 |
| Externalizing | 0.166145 | p = 0.140773 | 1.41E-01 |

**Supplemental Table 8.** SSRI exposure is associated with BOLD response to Negative-Neutral faces in girls.

| Brain Region | Beta value | p | p-FDR |
| --- | --- | --- | --- |
| Left Amygdala | **0.924693** | 0.000001 | 1.17E-09 |
| Right Amygdala | **0.519442** | 0.00036 | 7.53E-04 |
| Left Hippocampus | **0.828153** | 0.000001 | 1.17E-09 |
| Right Hippocampus | **0.595245** | 0.000002 | 6.83E-06 |
| Left Insula | **0.801484** | 0.00000002 | 1.26E-07 |
| Right Insula | **0.716672** | 0.0000005 | 2.50E-06 |
| Left Cuneus | 0.184558 | 0.175186 | 1.98E-01 |
| Right Cuneus | 0.206429 | 0.164478 | 1.98E-01 |
| Left Thalamus | **0.650476** | 0.000016 | 3.79E-05 |
| Right Thalamus | **0.47022** | 0.001145 | 2.14E-03 |
| Left Putamen | **0.678492** | 0.000004 | 1.29E-05 |
| Right Putamen | **0.654819** | 0.000012 | 3.38E-05 |
| Left rostral anterior cingulate cortex | 0.019179 | 0.891711 | 8.92E-01 |
| Right rostral anterior cingulate cortex | 0.024431 | 0.857061 | 8.92E-01 |
| Left Caudal ACC | **0.311557** | 0.029071 | 4.06E-02 |
| Right Caudal ACC | **0.45598** | 0.001587 | 2.68E-03 |
| Left Caudate | 0.249 | 0.072171 | 9.36E-02 |
| Right Caudate | **0.364782** | 0.009272 | 1.42E-02 |

**Supplemental Table 9.** SSRI exposure is not associated with symptoms in boys.

| Symptoms-Boys | Beta value | p-value | p-FDR |
| --- | --- | --- | --- |
| Anxiety | 0.217293 | 0.049117 | 0.1981121 |
| Depressive | -0.022278 | 0.852544 | 0.853 |
| Internalizing | 0.050892 | 0.658777 | 0.853 |
| Externalizing | -0.024 | 0.838148 | 0.853 |

**Supplemental Table 10.** SSRI exposure is not associated with BOLD response to Negative-Neutral faces in boys.

| Brain Region | Beta value | p | p-FDR |
| --- | --- | --- | --- |
| Left Amygdala | 0.177177 | 0.200178 | 0.257946 |
| Right Amygdala | 0.132437 | 0.380235 | 0.4029082 |
| Left Hippocampus | -0.151424 | 0.244835 | 0.294591 |
| Right Hippocampus | -0.185469 | 0.157728 | 0.2192126 |
| Left Insula | -0.227506 | 0.086006 | 0.1804574 |
| Right Insula | -0.229657 | 0.074964 | 0.1804574 |
| Left Cuneus | -0.272939 | 0.035148 | 0.1804574 |
| Right Cuneus | -0.220804 | 0.077806 | 0.1804574 |
| Left Thalamus | -0.143148 | 0.282498 | 0.3184177 |
| Right Thalamus | -0.21735 | 0.096066 | 0.1804574 |
| Left Putamen | -0.102482 | 0.42698 | 0.427331 |
| Right Putamen | -0.28074 | 0.027654 | 0.1804574 |
| Left rostral anterior cingulate cortex | -0.198635 | 0.140928 | 0.2192126 |
| Right rostral anterior cingulate cortex | -0.190629 | 0.153724 | 0.2192126 |
| Left Caudal ACC | -0.208265 | 0.095651 | 0.1804574 |
| Right Caudal ACC | -0.234467 | 0.07195 | 0.1804574 |
| Left Caudate | -0.212861 | 0.083446 | 0.1804574 |
| Right Caudate | -0.195206 | 0.099448 | 0.1804574 |

**Supplemental Table 11.**
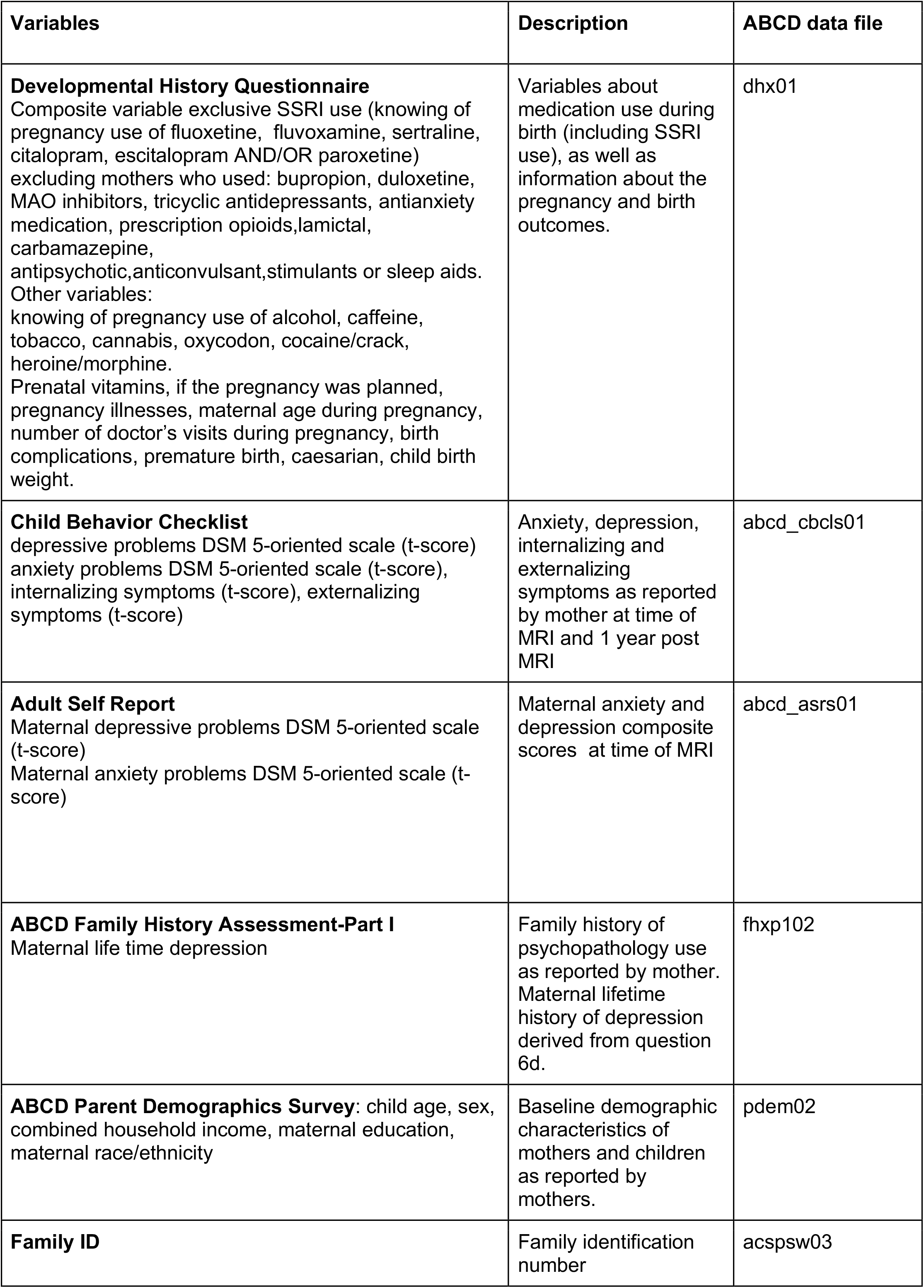
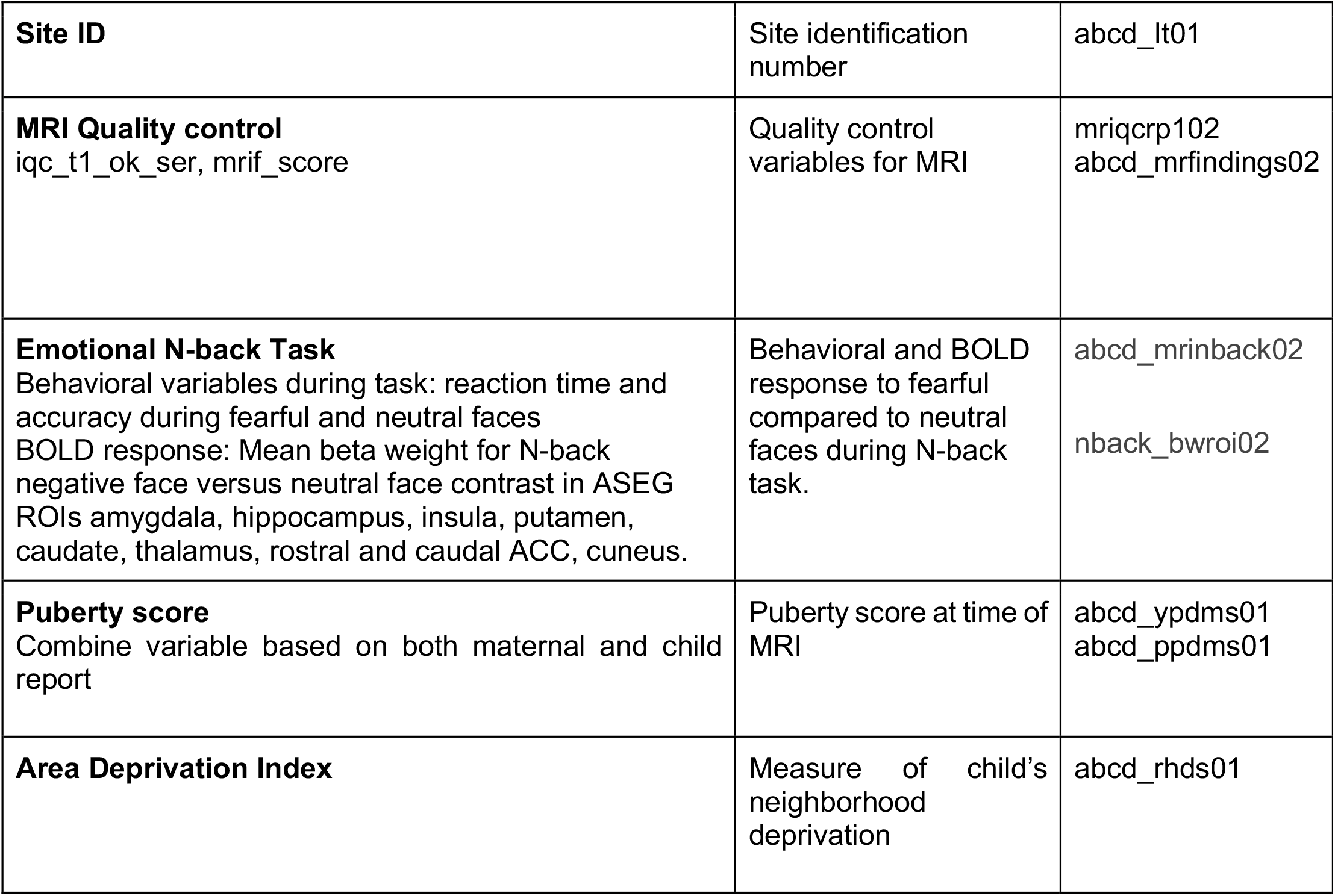
ABCD variables.

